# Cell-Mediated and Humoral Immune Response to 2-Dose SARS-CoV2 mRNA vaccination in Immunocompromised patient population

**DOI:** 10.1101/2021.07.21.21260921

**Authors:** Muthukumar Ramanathan, Kanagavel Murugesan, Lu M. Yang, Cristina Costales, Philip L. Bulterys, Joseph Schroers-Martin, Ash A. Alizadeh, Scott D. Boyd, Janice M. Brown, Kari C. Nadeau, Sruti S. Nadimpalli, Aileen X. Wang, Stephan Busque, Benjamin A. Pinsky, Niaz Banaei

**Affiliations:** Department of Pathology, Stanford University School of Medicine, Stanford, CA 94305 USA; Department of Medicine, Division of Oncology, Stanford University School of Medicine, Stanford, CA 94305 USA; Sean N. Parker Center for Allergy & Asthma Research, Stanford, CA 94305 USA; Department of Medicine, Division of Infectious Diseases and Geographic Medicine, Stanford University School of Medicine, Stanford, CA 94305 USA; Department of Medicine, Division of Pulmonary, Allergy & Critical Care Medicine, Stanford University School of Medicine, Stanford, CA 94305 USA; Department of Pediatrics, Division of Pediatric Infectious Diseases, Stanford University School of Medicine, Stanford CA 94305 USA; Department of Medicine, Division of Nephrology, Stanford University School of Medicine, Stanford, CA 94305 USA; Department of Surgery, Division of Abdominal Transplantation, Stanford University School of Medicine, Stanford, CA 94305 USA; Clinical Microbiology Laboratory, Stanford Health Care, Palo Alto, CA 94304 USA

## Abstract

Characterization of cell-mediated and humoral immune responses to SARS-CoV2 mRNA vaccine has implications for protective immunity in immunocompromised patients. However, studies have demonstrated poor humoral response to SARS-CoV2 mRNA vaccine in immunocompromised patients and data on cellular immune response are currently lacking. Here we compared immune response after 2-dose vaccination in 100 immunocompromised patients (solid organ transplant recipients, hematologic malignancy, autoimmune condition, and primary immunodeficiency) and 16 immunocompetent healthy healthcare workers. We find that 100% (CI=80.6-100%) of immunocompetent individuals show positive cell-mediated and humoral immune response post vaccination while only 50% (CI=40.4-59.6%) of immunocompromised patients show humoral immune response and 69% (CI=59.4-77.2%) have a positive cell-mediated immune response. 21% of immunocompromised patients have no humoral immune response or cell-mediated immune response and thus are likely vulnerable to SARS-CoV2 infection. Monitoring of immune response in immunocompromised populations, particularly in high-risk immunocompromised patients (solid organ transplant recipients, patients with severe autoimmunity, and those ≥50 years), with clinical IGRA and serological assay after vaccination may identify patients who may benefit from revaccination or prophylactic monoclonal antibody therapy to prevent COVID-19 in this patient population

---

One in 25 Americans are estimated to be immunocompromised ^1^. Characterization of cell-mediated and humoral immune responses to SARS-CoV2 vaccine has implications for protective immunity in this vulnerable population. However, studies have demonstrated poor humoral response to SARS-CoV2 mRNA vaccine in immunocompromised patients ^2^ and data on cellular immune response are currently lacking. Hence, the nature of cellular immune response in immunocompromised patients after standard 2 dose mRNA vaccination is unclear compared to immunocompetent individuals, in whom the vaccine trials demonstrated robust cellular and humoral immune responses.

Here, we compared cell-mediated and humoral immune responses to SARS-CoV-2 mRNA vaccination in immunocompromised patients and immunocompetent controls. Clinically orderable laboratory-developed interferon-γ release assay (IGRA) was performed to measure and qualitatively interpret T-cell response to SARS-CoV2 antigens. Humoral response was evaluated using a commercial ELISA assay measuring IgG for the SARS-CoV-2 spike S1 domain. Immunocompetent controls (16 healthy healthcare workers, median age 42 [IQR 36-47]; 38% men) provided informed consent for post-vaccination immune response testing after (median 17 days [IQR 15-18]) the second dose of BNT162b2 mRNA vaccine. Immunocompromised patients (100 patients, median age 56 [IQR 36-68]; 54% men) were tested by their providers after (median 50 days [IQR 45-79]) the second dose of Moderna or Pfizer/BioNTech mRNA vaccine. Immunocompromised patients comprised four subgroups - solid organ transplant (n=21), hematologic malignancy (n=38), autoimmune condition treated with antimetabolite drugs or biologics (n=17) or primary immunodeficiency (n=24).

100% (95% confidence interval (CI) = 80.6–100%) of immunocompetent controls had a positive IGRA qualitative result compared to 69% (CI=59.4-77.2%) in all immunocompromised patients (Figure 1A). However, there was no significant difference in median interferon-γ response in controls when compared with immunocompromised patients (1.29 IU/mL vs 1.23 IU/mL; p=0.2) (Figure 1B). All 16 (100%, CI=80.6-100%) controls had a positive qualitative antibody result compared to 50% (CI=40.4-59.6%) positivity rate in immunocompromised patients (Figure 1C). Immunocompetent controls had higher median antibody IgG levels compared to immunocompromised patients (12 vs 2.42 OD ratio, p<0.0001) (Figure 1D). In 29% of immunocompromised patients, cell-mediated response was the only positive immune response to the vaccine compared with 10% for whom IgG was the only positive immune response (Supplementary Figure 1).

**Fig. 1.**
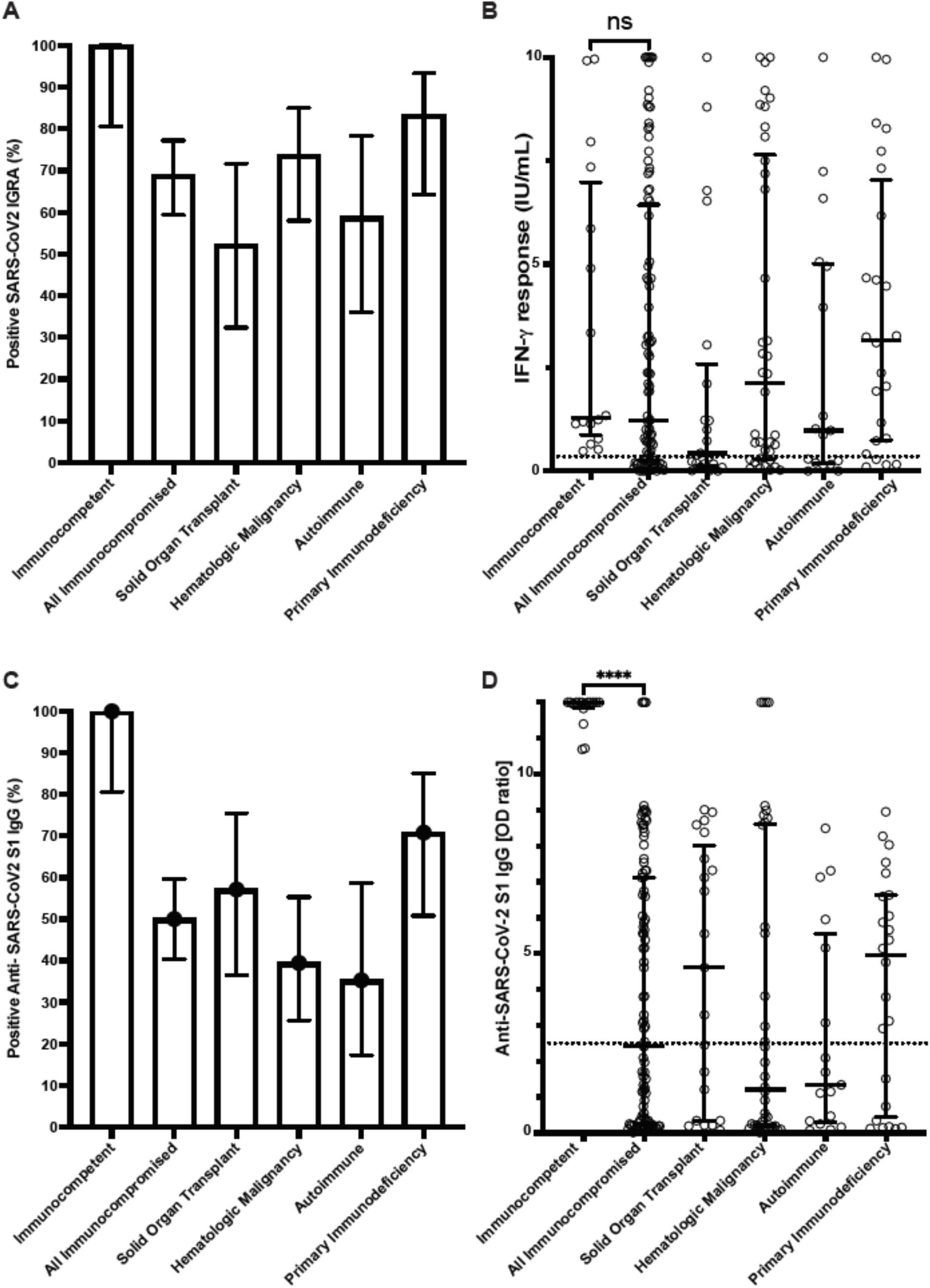
Cellular and humoral immune responses to SARS-CoV-2 mRNA vaccine in immunocompetent individuals and immunocompromised patients determined with clinical assays. (A) Percentage of patients with positive SARS-CoV2 interferon-γ release assay (IGRA) result. (B) Interferon-γ response from IGRA in immunocompetent individuals (n=16) and immunocompromised patients (n=100) (C) Percentage of patients with positive Anti-SARS-CoV2 S1 IgG ELISA result. (D) Anti-SARS-CoV2 S1 IgG optical density (OD) ratio from ELISA performed in immunocompetent and immunocompromised patients. Results for immunocompromised subgroups are also shown. Dotted lines represent the assay cutoffs (0.35 IU/mL for IGRA and 2.5 optical density ratio for IgG). Long solid lines show median, short solid lines show 95% confidence interval. ns, not significant and ****, p<0.0001 by Mann-Whitney U test.

Comparing cell-mediated and humoral immune responses among all immunocompromised patients, no significant difference was noted based on mRNA vaccine manufacturer (Moderna vs Pfizer/BioNTech) or sex (Supplementary Figure 2). However, immunocompromised patients below the age of 50 had 5-fold higher median IgG levels than patients age 50 or older (5.90 vs 1.21 OD ratio; p<0.001). No significant difference was noted in cell-mediated response based on age among immunocompromised patients (Supplementary Figure 3).

This study demonstrates that immunocompromised patients have a higher rate of positive cell-mediated response (69%, CI=59.4-77.2%) than humoral response (50%, CI=40.4%-59.6%) to SARS-CoV-2 mRNA vaccine and interferon-γ levels are comparable between immunocompetent controls and immunocompromised patients. IgG levels, however, were significantly lower in the latter group. The absence of either cell-mediated or humoral immune response was highest in solid organ transplant recipients (38.1%) and patients with autoimmune disorders (35.3%) (Supplementary Figure 4).

Prior studies in patients with influenza have demonstrated that cell-mediated immune response to influenza virus in the absence of humoral response correlates with protection against symptomatic influenza^4^. Further studies are required to confirm protection against COVID-19 in vaccinated, immunocompromised patients with cell-mediated immune response alone. In contrast to immunocompetent healthcare workers, in whom 100% had positive cellular and humoral immune response, this study revealed that a significant fraction (21%) of immunocompromised patients mount no cellular or humoral response after vaccination and likely remain susceptible to infection with ancestral strains or emerging variants of SARS-CoV2. Monitoring of immune response in immunocompromised populations, particularly in high-risk immunocompromised patients (solid organ transplant recipients, patients with severe autoimmunity, and those ≥50 years), with clinical IGRA and serological assay after vaccination may identify patients who may benefit from revaccination^5^ or prophylactic monoclonal antibody therapy to prevent COVID-19 in this patient population.

## Data Availability

All data relevant to the study are included in the article. De-identified data of this study could be shared upon request.

---

**Fig. S1.**
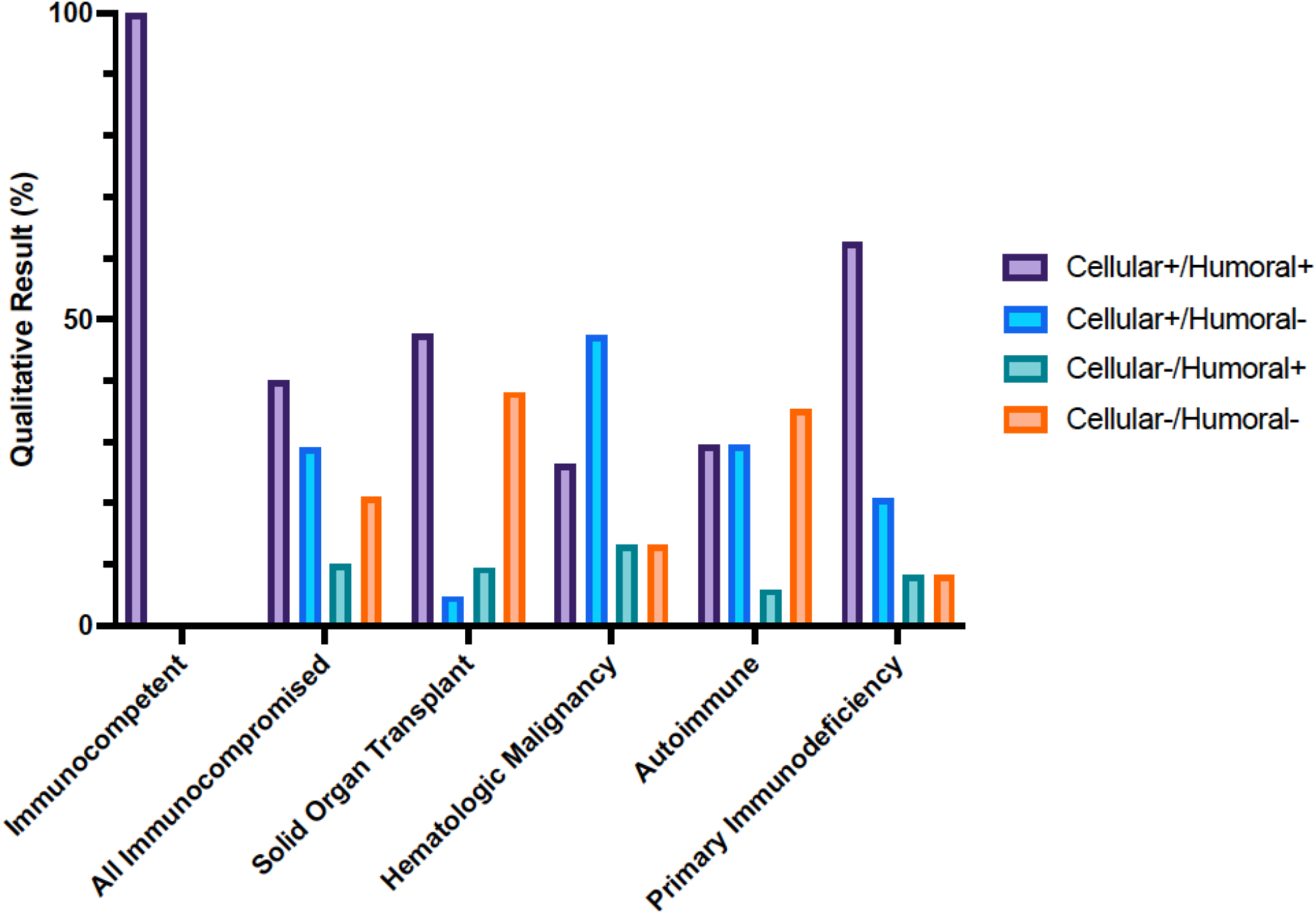
Cellular and humoral immune responses to SARS-CoV-2 mRNA vaccine in immunocompetent and immunocompromised patients determined with clinical assays. Percentage of immunocompetent and immunocompromised patients with positive or negative SARS-CoV2 interferon-γ release assay and Anti-SARS-CoV-2 S1 IgG ELISA result after 2 doses of SARS-CoV2 mRNA vaccination.

**Fig. S2.**
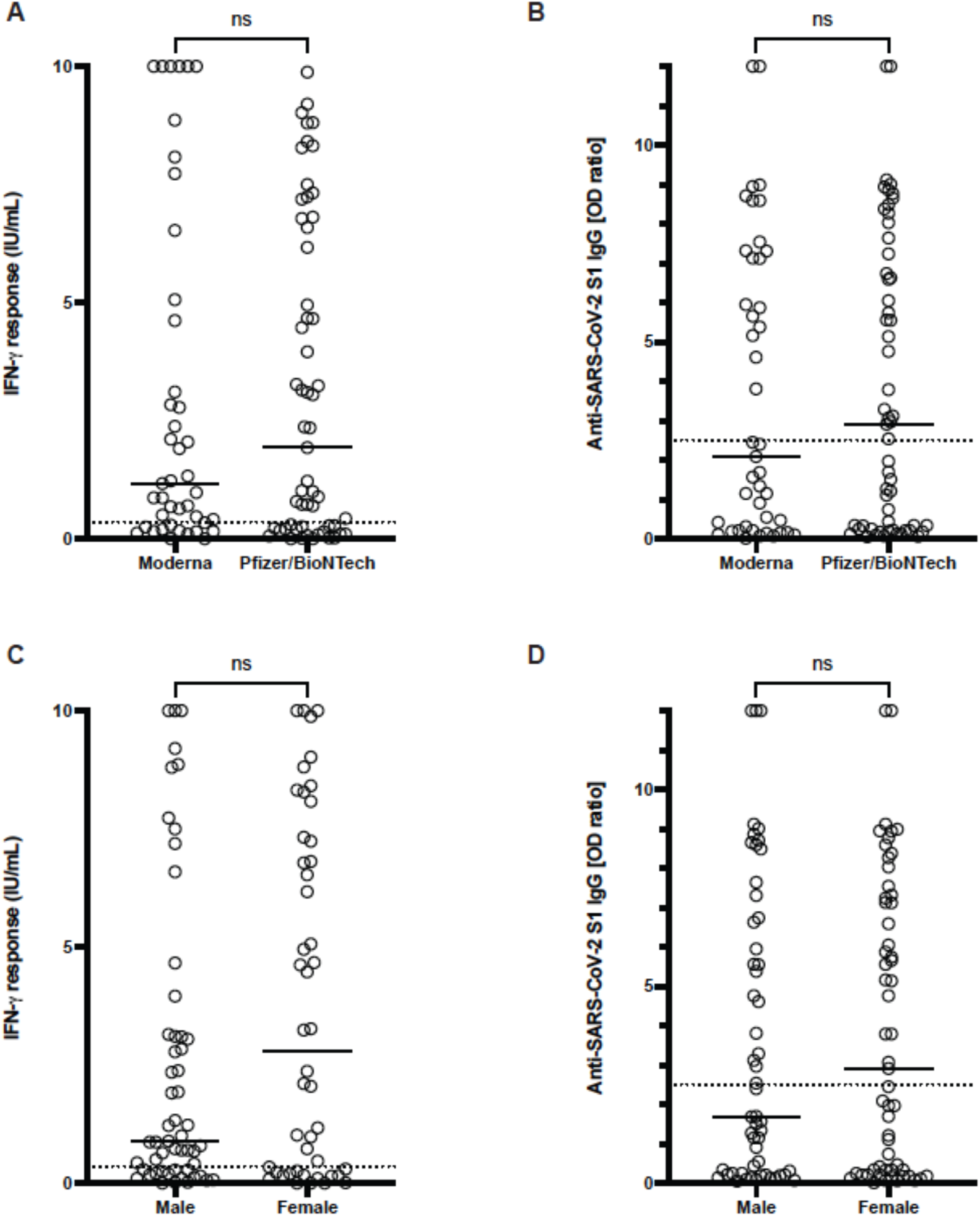
Cellular and humoral immune responses to SARS-CoV-2 mRNA vaccine in immunocompromised patients based on mRNA vaccine manufacturer and sex. (A and C) Interferon-γ response with interferon-γ release assay. (B and D) IgG levels with Anti-SARS-CoV-2 S1 IgG ELISA. Tests were performed in immunocompromised patients (n=100) who received 2 doses of Moderna (n=43) or Pfizer/BioNTech (n=57) SARS-COV2 mRNA vaccine. Dotted lines represent the assay cutoffs (0.35 IU/mL for IGRA and 2.5 optical density ratio for IgG). Solid lines show median. ns, not significant by Mann-Whitney U test.

**Fig. S3.**
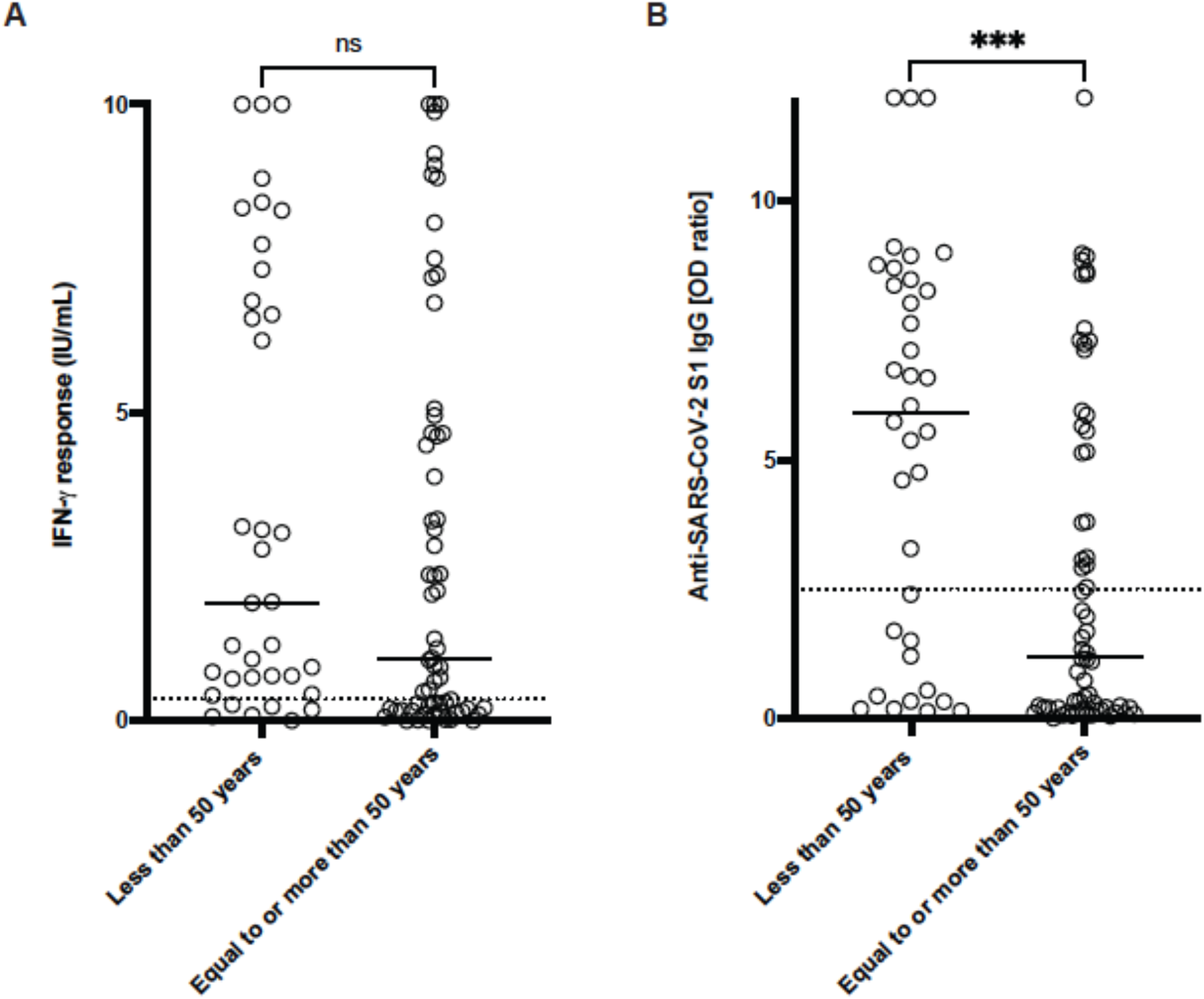
Cellular and humoral immune responses to SARS-CoV-2 mRNA vaccine in immunocompromised patients based on age. (A) Interferon-γ response with interferon-γ release assay. (B) IgG levels with Anti-SARS-CoV-2 S1 IgG ELISA. Tests were performed in immunocompromised patients (n=100) comparing median age below 50 (n=36) with 50 or above (n=64). Dotted lines represent the assay cutoffs (0.35 IU/mL for IGRA and 2.5 optical density ratio for IgG). Solid lines show median. ns, not significant and ***, p<0.001 by Mann-Whitney U test.

**Fig. S4.**
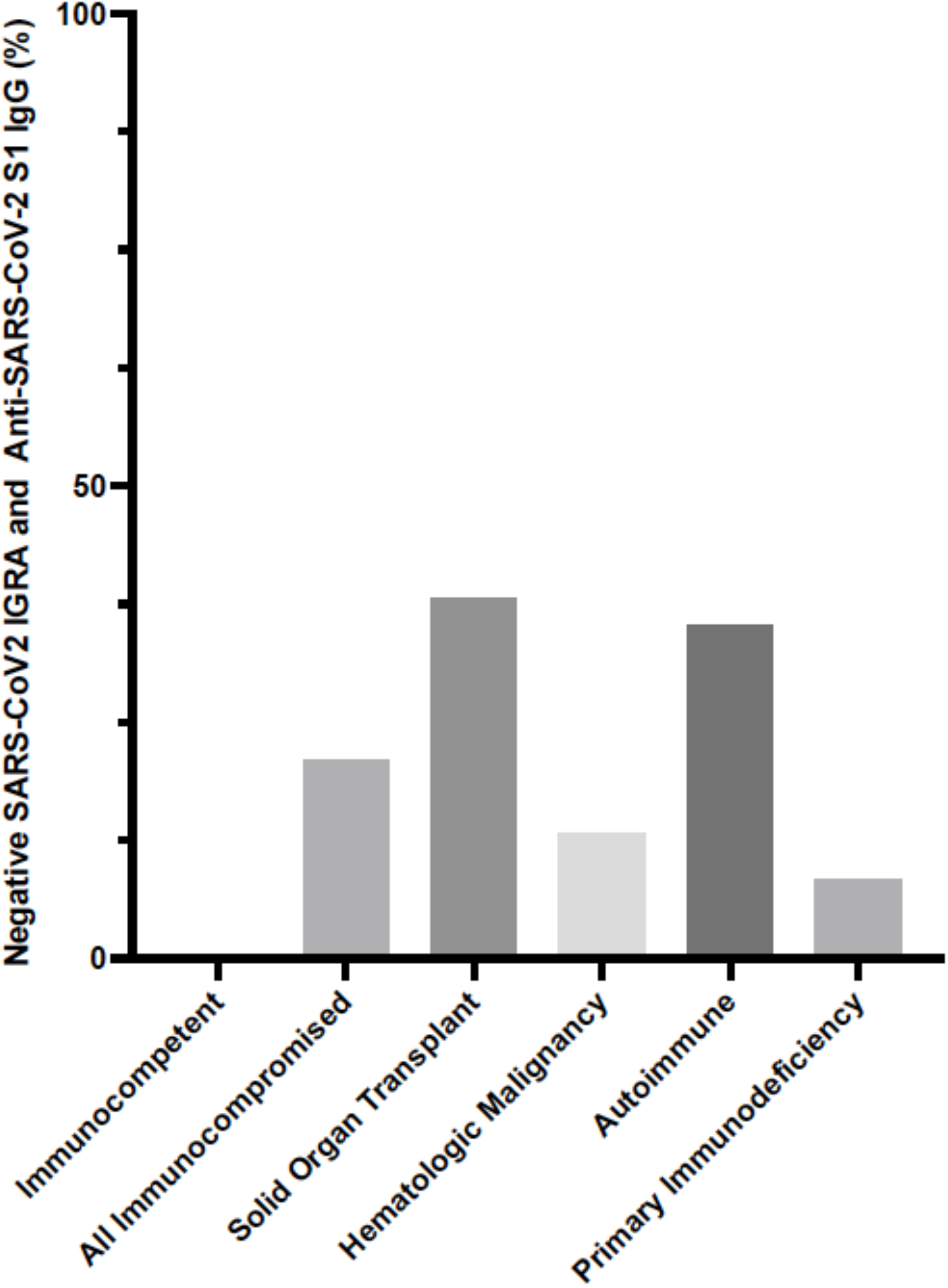
Negative cellular and humoral immune responses to SARS-CoV-2 mRNA vaccine in immunocompetent and immunocompromised patients determined with clinical assays. Percentage of patients with negative SARS-CoV2 Interferon-γ release assay and negative Anti-SARS-CoV2 S1 IgG ELISA results after 2 doses of SARS-CoV2 mRNA vaccination.

## Material and Methods

### Ethics

This study was approved by the Stanford University Institutional Review Board (IRB-60171 and IRB-57519). Healthcare worker participants provided informed consent to donate blood samples via venipuncture after second dose of vaccination with BNT162b2. IGRA and IgG ELISA were ordered in immunocompromised patients by their providers as part of clinical testing. Retrospective chart review of immunocompromised patients was performed to obtained clinical data.

### Interferon Gamma Release Assay (IGRA)

SARS-CoV-2 IGRA was performed in the Stanford Health Care Clinical Microbiology Laboratory as described previously ^1^. Freshly collected blood in lithium heparin tube was transferred to BD vacutainer no additive tubes at 1mL per tube. One tube was left unstimulated (nil), one tube was stimulated with SARS-CoV-2 antigen peptide megapool (Miltenyi Biotec, Bergisch Gladbach, Germany) at 2.2 mmol/mL, and one tube was stimulated with Phytohemagglutinin PHA-P Mitogen (Sigma, St. Louis, MO) at 50µL/mL. The blood samples were mixed gently and incubated at 37 °C for 20 to 24 h. The interferon-γ concentration was measured in the plasma portion with an automated enzyme-linked immunosorbent assay (ELISA) instrument (DSX; Dynex Technologies, Chantilly, VA) using the QuantiFERON-TB ELISA kit (Qiagen, Germantown, MD). Interferon-γ response was defined as positive if antigen-nil ≥0.35 IU/mL; negative if antigen-nil <0.35 and mitogen-nil ≥0.5 IU/mL, and indeterminate if nil >8 IU/mL or antigen-nil <0.35 and mitogen-nil <0.5 IU/mL. All values higher than 10 IU/mL were adjusted to 10 IU/mL to reflect the linear range of assay.

### Enzyme-linked immunosorbent assay (ELISA)

SARS-CoV-2 Spike S1 IgG serology was performed by semi-quantitative ELISA (EUROIMMUN, Lübeck, Germany) according to the manufacturer’s instructions. Briefly, 1:100 diluted serum samples from vaccinees were added to the antigen pre-coated microplate followed by the addition of anti-human IgG conjugate, chromogenic substrate and stopping solution. The optical density (OD) was measured at 450nm. Results were evaluated by calculating the ratio of the optical density (OD) of the sample over the OD of a calibrator provided by the manufacturer. Assay cutoff values were defined as follows: <2.5 negative, ≥2.5 positive ^2^. All values higher than OD 12 were adjusted to 12 to reflect the linear range of assay.

### Statistics

GraphPad Prism version 9.0 (GraphPad Software, San Diego, California, USA) software was used to visualize data and Mann-Whitney U test was performed to compare cellular and humoral response in immunocompetent and immunocompromised patients.

## Acknowledgements

We thank all study participants, providers and nursing staff caring for immunocompromised patients evaluated in this study.

---

**Table S1:** Demographic and clinical characteristics of study participants (Immunocompetent and Immunocompromised)

| **Immunocompetent (n=16)** |  |
| --- | --- |
| Male | 6 (37.5%) |
| Female | 10 (62.5%) |
| Age (Median, IQR) | 42 (36-47) |
| **Vaccine** |  |
| A) Pfizer/BioNTech | 16(100%) |
| B) Moderna | 0(0%) |
| **Immune Response** |  |
| 1) Cellular Positive & Humoral Positive | 16(100%) |
| 2) Cellular Positive & Humoral Negative | 0(0%) |
| 3) Cellular Negative & Humoral Positive | 0(0%) |
| 4) Cellular Negative & Humoral Negative | 0(0%) |

| **Immunocompromised (n=100)** |  |
| --- | --- |
| Male | 54 (54%) |
| Female | 46 (46%) |
| Age (Median, IQR) | 56 (36-68) |
| **Vaccine** |  |
| A) Pfizer/BioNTech | 57(57%) |
| B) Moderna | 43(43%) |
| **Immune Response** |  |
| 1) Cellular Positive & Humoral Positive | 40(40%) |
| 2) Cellular Positive & Humoral Negative | 29(29%) |
| 3) Cellular Negative & Humoral Positive | 10(10%) |
| 4) Cellular Negative & Humoral Negative | 21(21%) |

| **Solid Organ Transplant (n=21)** |  |
| --- | --- |
| Male | 10 (47.6%) |
| Female | 11 (52.3%) |
| Age (Median, IQR) | 39 (18-62) |
| **Vaccine** |  |
| A) Pfizer/BioNTech | 13(61.9%) |
| B) Moderna | 8(38.1%) |
| **Transplants** |  |
| Renal Transplant | 15(71.4%) |
| Liver Transplant | 5(23.8%) |
| Heart Transplant | 1(4.8%) |
| **Immune Response** |  |
| 1) Cellular Positive & Humoral Positive | 10(47.6%) |
| 2) Cellular Positive & Humoral Negative | 1(4.8%) |
| 3) Cellular Negative & Humoral Positive | 2(9.5%) |
| 4) Cellular Negative & Humoral Negative | 8(38.1%) |

| **Hematological Malignancy (n=38)** |  |
| --- | --- |
| Male | 28 (73.6%) |
| Female | 10 (26.3%) |
| Age (Median, IQR) | 60 (40-70) |
| Vaccine |  |
| A) Pfizer/BioNTech | 21(55.3%) |
| B) Moderna | 17(44.7%) |
| Immune Response |  |
| 1) Cellular Positive & Humoral Positive | 10(26.3%) |
| 2) Cellular Positive & Humoral Negative | 18(47.4%) |
| 3) Cellular Negative & Humoral Positive | 5(13.2%) |
| 4) Cellular Negative & Humoral Negative | 5(13.2%) |

| **Autoimmune condition (n=17)** |  |
| --- | --- |
| Male | 8 (47.1%) |
| Female | 9 (52.9%) |
| Age (Median, IQR) | 67 (57-71) |
| Vaccine |  |
| A) Pfizer/BioNTech | 7(41.2%) |
| B) Moderna | 10(58.8%) |
| Autoimmune condition |  |
| Rheumatoid Arthritis | 9 (52.9%) |
| Other Autoimmune (MS, CIDP etc) | 8 (47.1%) |
| Immune Response |  |
| 1) Cellular Positive & Humoral Positive | 5(29.4%) |
| 2) Cellular Positive & Humoral Negative | 5(29.4%) |
| 3) Cellular Negative & Humoral Positive | 1(5.9%) |
| 4) Cellular Negative & Humoral Negative | 6(35.3%) |

| **Primary Immunodeficiency (n=24)** |  |
| --- | --- |
| Male | 8 (33.3%) |
| Female | 16 (66.7%) |
| Age (Median, IQR) | 48 (34-65) |
| Vaccine |  |
| A) Pfizer/BioNTech | 16(66.7%) |
| B) Moderna | 8(33.3%) |
| Immunodeficiency |  |
| CVID | 13 (54.2%) |
| Others (Hypogammaglobulinemia etc) | 11 (45.8%) |
| Immune Response |  |
| 1) Cellular Positive & Humoral Positive | 15(62.5%) |
| 2) Cellular Positive & Humoral Negative | 5(20.8%) |
| 3) Cellular Negative & Humoral Positive | 2(8.3%) |
| 4) Cellular Negative & Humoral Negative | 2(8.3%) |

